# Public Health Interventions for Fractional Optimal Control of Buruli Ulcer

**DOI:** 10.1101/2024.09.05.24313151

**Authors:** Solomon Nortey, Ernest Akorly, Mark Dadzie, Stephen E. Moore

## Abstract

Buruli Ulcer, a devastating skin disease caused by *Mycobacterium Ulcerans*, poses considerable public health challenges in endemic areas. This article focuses on the use of fractional optimal control theory to prevent the spread of Buruli ulcers via integrated public health interventions. We formulated a mathematical model using the Atangana-Baleanu-Caputo fractional order derivative operator. We investigated the model’s existence and uniqueness and presented numerical simulations using the predict-evaluate-correct-evaluate (PECE) method of Adam-Bashforth Moulton. We also study the fractional optimal control problem (FOCP) to minimize the spread of the disease in the endemic regions. We employ the Fractional Pontryagin’s Maximum Principle (FPMP) and implement the forward-backward method to determine the extremals of the problem. Four control strategies were implemented: promoting health education on the use of protective clothing, enhancing vaccination rates, improving treatment protocols for infected individuals, and spraying insecticides to reduce water-bug populations. After examining the optimal control dynamics of the Buruli ulcer transmission model via multiple simulations with and without control, we discover that there is a substantial decrease in the population of infected humans and the water-bug population. Hence we conclude that the best strategy to implement is by applying all the control strategies suggested.

## 1 Introduction

Buruli ulcer is a mysterious necrotizing tropical skin disease which is found mainly in the tropical regions with high cases recorded in Africa, America, Asia and Western Pacific [51, 36, 27, 58]. Amongst the 20 countries in Africa that cases have been recorded, Ghana recorded over 11,000 cases, Cote d’Ivoire recorded 21,000 and Togo reported over 2,000 cases[51]. While previously considered a childhood disease, statistical analysis shows that over 25% of affected individuals are over 50 years old [7].

The disease-causing organism belongs to the same family of bacteria that causes leprosy and tuberculosis, presenting a possibility for collaboration between the two disease programs [29]. Whereas the disease is known to be linked to contaminated water, according to the authors of [35] the mode of transmission to humans is still unclear, which makes it difficult to propose control interventions[50]. While the mode of transmission remains unresolved, once the causative bacterial, Mycobacterium Ulcerans enters the skin through direct injury or bites from insects like water bugs or mosquitoes as hypothesized in literature, it releases the Mycolactone Toxin [20, 34]. This toxin is responsible for the immunosuppression, cytotoxicity, modulation of host cell function and ultimately, the proliferation of Mycobacterium Ulcerans [14, 22]. Buruli Ulcer starts as nodule with no pains in patients but develops into painless ulcerating wound with weakened edges[16, 24]. Due to its painlessness [18], patients who mostly live in rural areas report late for treatment due to the reliance on traditional medication [49] and by which at the time of reporting, the ulcerating wound might have reached a severe stage which may lead to amputation of body part(s) or weeks to months of hospitalization. The cost and long duration of treatment coupled with its associated stigma and economic hardships that comes to bear on patients and their immediate aids is very alarming [9]. However, although clinical treatment is the ultimate effective preventive measure, one of the effective control measures for Buruli Ulcer is promoting education on the relevance of early detection through targeted programs and campaigns can increase public awareness of the availability and of clinical interventions[56] thereby reducing the disease’s stress.

Although several mathematical models on BU are found in literature, few of these models provide insight into the understanding of the dynamics of the transmission of the disease, efficient and effective control measures and the use of the model to predict a suitable prevention technique. The authors in [29] developed a non-linear mathematical model to examine the optimal control of transmission dynamics of Mycobacterium Ulcerans and obtained qualitative results using theories of stability of differential equations, optimal control and computer simulations. The authors employed two optimal control conditions, that is environmental and health education to people for prevention and to apply water and environmental purification rate. Based on the numerical results obtained, the authors established that, application of optimal control leads to the decrease of the number of infected waterbugs and also decreases the number of human infected by MU. However the authors concluded that, in order to reduce the spread of MU infection,the application of optimal control on environmental and health education in human must be used for prevention. Although [15] and [36] used somewhat different optimal control conditions, they yielded similar results and conclusions to those found in [29]. In [21], the authors used fractional and integer derivatives to study the dynamics of the transmission of Buruli ulcer. They established that, in quantitative sense, the fractional model used in the study presented knowledge of the history as compared to the classical model. Nevertheless, the authors admitted that all results obtained are limited to fractional derivatives in the Caputo sense and expressed uncertainty in the possible results of using other fractional derivatives such as the Caputo Fabrizio or Atangana–Baleanu. Again the authors did not incorporate any optimal control conditions but rather maintained it as a constant. However, in this study, we formulated a mathematical model to prevent the spread of Buruli Ulcer using the Atangana-Baleanu-Caputo fractional order derivative operator as well as public health interventions.

The rest of the article is organized as follows: the preliminaries of fractional calculus are introduce in section 2, the fractional BU model is derived in section 3, the analysis of the BU model is discussed in section 4, the numerical simulation of the BU model is discussed in section 5, section 6 describe the fractional optimal control problem, the numerical simulation and discussion of optimal control problem is presented in section 7 and finally we conclude in section 8.

## 2 Mathematical Preliminary on Fractional Calculus

We present important definitions and Lemma necessary for the development and analysis of the fractional model.

### Definition 2.1

([39, 42]). *The Atangana-Baleanu fractional derivative in the Caputo sense (ABC) with order α* ∈ (0, 1] *and lower limit zero for a function g* ∈ *H*^1^(0, *T*) *is defined by*

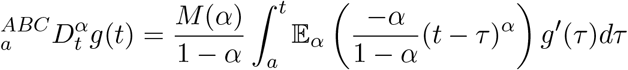

*where* 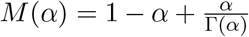 *is the normalization function satisfying M* (0) = *M* (1) = 1, *and* 𝔼_*α*_ *is the Mittag-Leffler function expressed as*

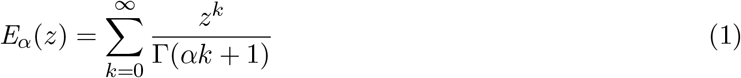

### Definition 2 .2

([19]). *The associated ABC f ractional integral i s defined by*

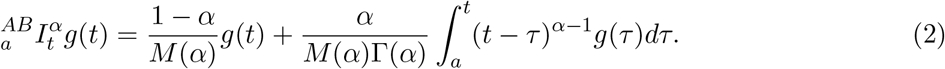

### Definition 2.3

([39, 42]). *The Laplace transformation of the equation above is expressed as*

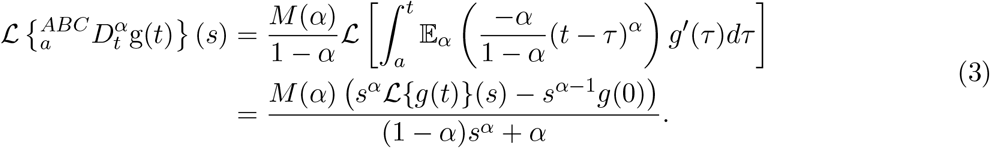

### Lemma 2.4

([6, 12]). *If α* ∈ (0, 1] *and h*(*t*) ∈ *C*[0, 1]. *Then the solution to*

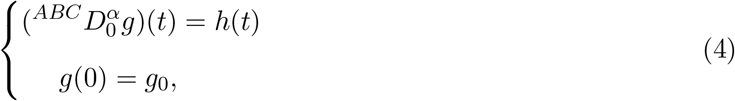

*is given by*

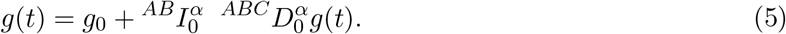

## 3 Fractional Order Model Derivation

Using systems of nonlinear differential e quations, we b uild u p a c ompartmental m odel f or the transmission dynamics of Buruli Ulcer. The model includes two population, that is human and water-bug populations.The total human population is given by *N*_*H*_(*t*) is subdivided into five classes; Susceptible *S*_*H*_(*t*), Vaccinated, *V*_*H*_(*t*), Exposed *E*_*H*_(*t*), Infected *I*_*H*_(*t*) and Recovered *R*_*H*_(*t*). Hence the dynamics of the total human population is

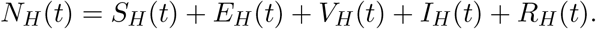

The second population which is the water-bug has a total population *N*_*w*_(*t*) which is subdivided into Susceptible water-bug *S*_*w*_(*t*) and Infected water-bug class *I*_*w*_(*t*). Hence the dynamic of the total water-bugs population is

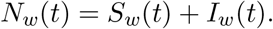

We consider Λ_*H*_ to be the recruitment rate into the susceptible human class. We assume that susceptible humans get infected when bitten by infected water-bugs, hence the force of infection is given by *ρβ*_1_*S*_*H*_*I*_*w*_ where *ρ* is the biting rate and *β*_1_ is the transmission probability rate. We also assume that the vaccine is imperfect, hence vaccinated individuals can be infected when bitten by infected water-bugs at a reduce rate of (1 − *σ*). The parameters *τ* and *λ* are the rate at which susceptible humans are vaccinated and the vaccine wane rate respectively. *µ*_*H*_ is the natural mortality rate that occurs in the human population. Hence the dynamics of both the susceptible and vaccinated class is

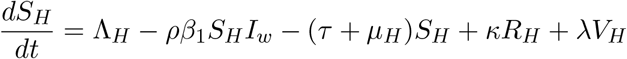

and

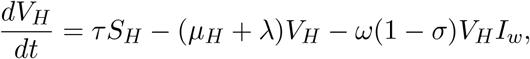

respectively. The exposed class increases by *ρβ*_1_*S*_*H*_*I*_*w*_ and *ω*(1 − *σ*)*V*_*H*_*I*_*w*_ while it decreases by (*µ*_*H*_ + *ε*) where *ε* is the rate at which exposed class become infected. Hence the dynamics of the exposed class is

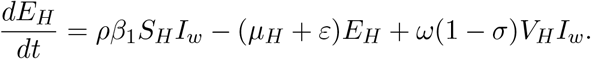

The infected human class increases at the rate by which individuals leave the exposed class *εE*_*H*_ and decreases by (*µ*_*H*_ + *δ* + *γ*). Therefore the dynamics of the infected human class is

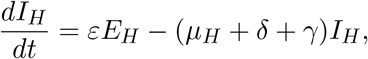

where the *δ* and *γ* are death induce by the disease and recovery rate respectively. The recovery class increases by the rate *τ* and decreases by (*µ*_*H*_ + *κ*) where *κ* is the rate at which recovered individuals become susceptible again. Hence the dynamics of the recovered class is

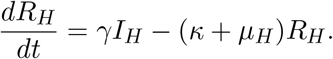

The susceptible water-bug becomes infected by biting an infected human and coming into contact with the Mycobacterium Ulceran, hence the force of infection is given by *ρβ*_2_*S*_*w*_*I*_*H*_. The dynamics of the susceptible water-bug class is

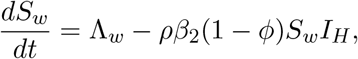

where *ϕ* is the rate at which susceptible water-bugs are infected by coming into contact with the Mycobacterium Ulceran environment. Lastly the dynamics of the infected water-bugs is given by

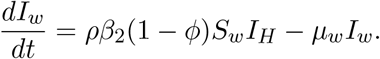

The following assumptions were made in order to derive the Buruli Ulcer model[36]. The transmission dynamics consist of two populations, human and water-bug population, the pathogen is transferred from waterbugs to humans and vice versa, Distinct recruitment and death rates, Imperfect Vaccination: Hence vaccinated, individuals can be infected when bitten by infected water-bugs at reduce rate of (1 − *σ*),potential reinfection of recovered individuals, the population of water-bugs is higher than that of humans and the rate at which water-bugs come into contact with Mycobacterium Ulcerans in their environment is *ϕ*.

In Fig (3) and system (6), the schematic diagram and equations are described respectively.

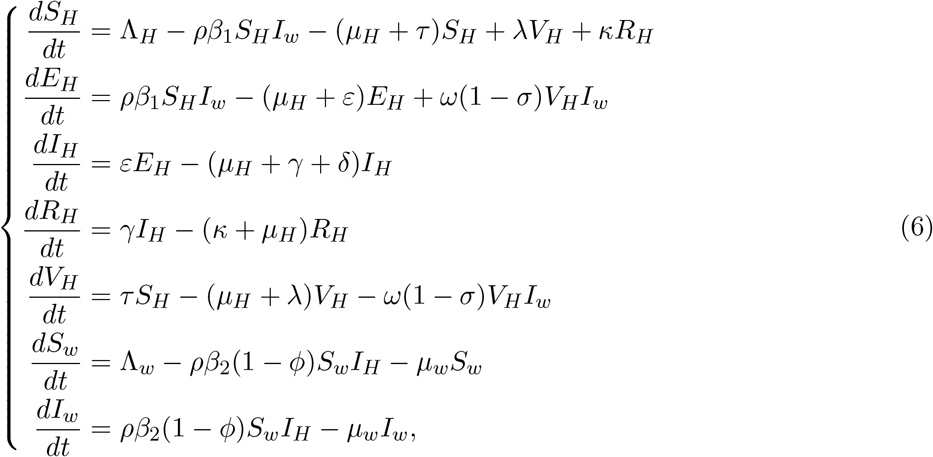

with positive initial conditions *S*_*H*_(0) *>* 0, *V*_*H*_(0) ≥ 0, *E*_*H*_(0) ≥ 0, *I*_*H*_(0) ≥ 0, *R*_*H*_(0) ≥ 0, *S*_*w*_(0) *>* 0 and*I*_*w*_(0) ≥ 0.

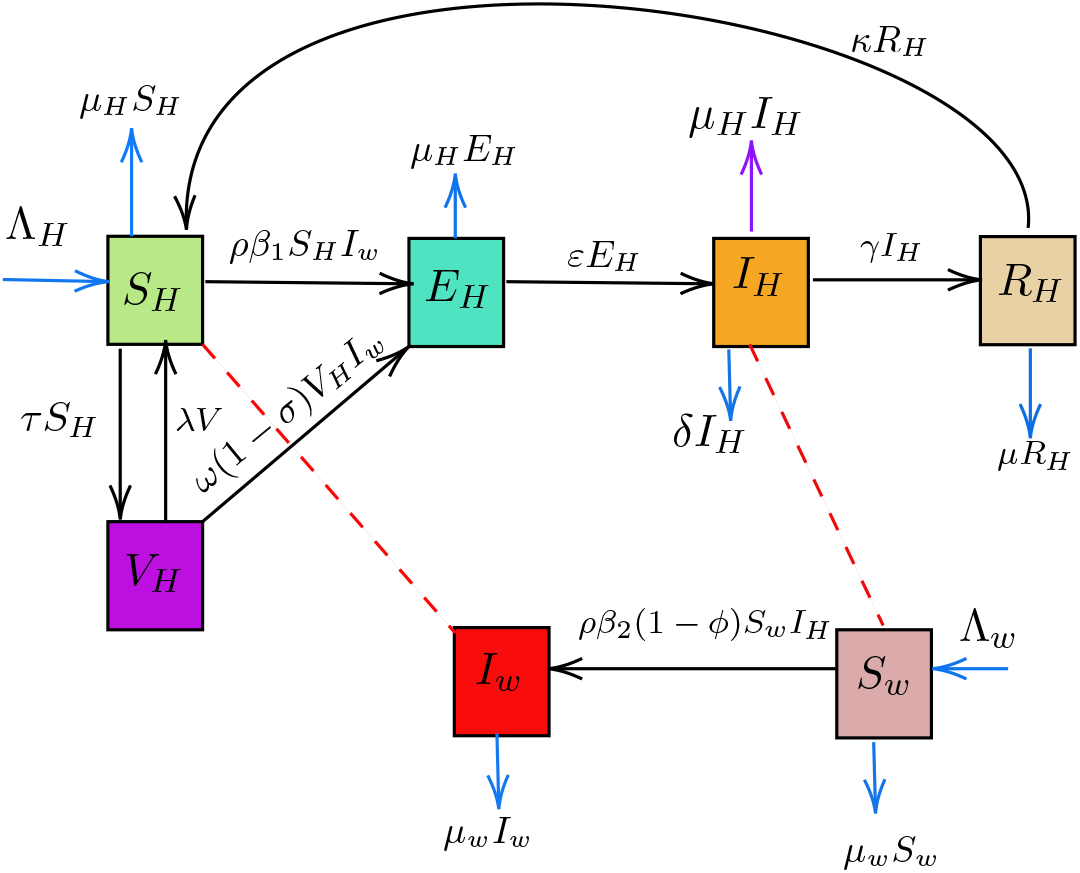

The compartmental model for Buruli Ulcer

We now formulate the Atangana-Baleanu Caputo (ABC) fractional order derivative form of the equation (6) as

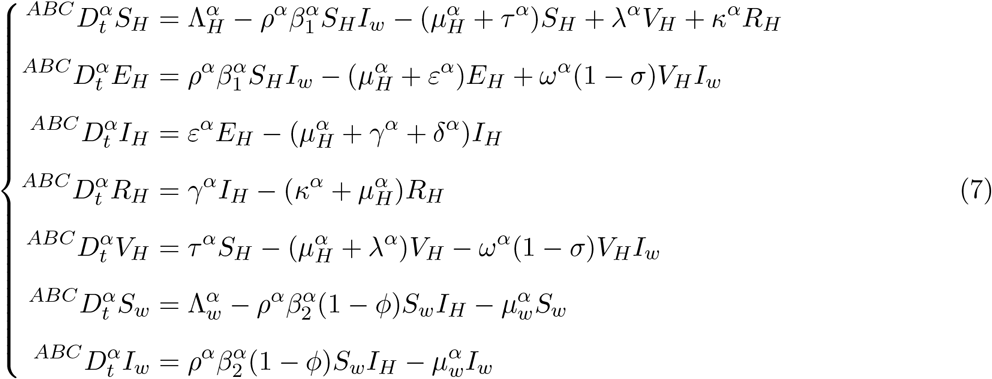

with the initial conditions *S*_*H*_(0) *>* 0, *V*_*H*_(0) ≥ 0, *E*_*H*_(0) ≥ 0, *I*_*H*_(0) ≥ 0, *R*_*H*_(0) ≥ 0, *S*_*w*_(0) *>* 0 and *I*_*w*_(0) ≥ 0, where 0 ≤ *t < T* and 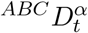 denotes the Atangana-Baleanu Caputo fractional derivative of order *α* ∈ (0, 1]. The parameters and variables of the model (7) are described in detail below, see Table 1.

**Table 1:** Parameter and Variable Descriptions.

| Parameter | Description |
| --- | --- |
| $\beta_1$ | The transmission probability of infected water-bugs. |
| $\rho$ | The biting rate of infected water-bugs on susceptible individuals. |
| $\mu_H$ | The natural mortality rate of the human population. |
| $\varepsilon$ | Represent the rate at which exposed class become infected. |
| $\gamma$ | The recovery rate. |
| $\Lambda_H$ | Birth rate of susceptible humans. |
| $\Lambda_w$ | The recruitment rate of susceptible water-bugs. |
| $\beta_2$ | The transmission probability of infected humans . |
| $\kappa$ | The rate at which recovered individuals become susceptible. |
| $\delta$ | Represent the induced death rate by the disease. |
| $\sigma$ | Efficacy of the vaccine. |
| $\phi$ | The proportion of Mycobacterium Ulceran infecting water-bugs. |
| $\tau$ | Vaccination rate. |
| $\lambda$ | Vaccine wane rate. |

## 4 Mathematical Analysis of the Model

In this section, we consider the qualitative aspects of the model (7). We present the proof of existence and uniqueness of the solution by means of fixed point iteration technique in specific norm.

### 4.1 Existence And Uniqueness Of Solution

**Theorem 4.2**. *[41] Suppose that F* (*X*) *is a Banach space of real-valued continuous functions defined on the interval X* = [0, *T* ] *with the sup norm, and let G* = *F* (*X*)*×F* (*X*)*×F* (*X*)*×F* (*X*)*×F* (*X*)*×F* (*X*) *with the norm* ∥(*S*_*H*_, *E*_*H*_, *I*_*H*_, *R*_*H*_, *V*_*H*_, *S*_*w*_, *I*_*w*_)∥ = ∥*S*_*H*_∥ + ∥*E*_*H*_∥ + ∥*I*_*H*_∥ + ∥*R*_*H*_∥ + ∥*V*_*H*_∥ + ∥*S*_*w*_∥ + ∥*I*_*w*_∥,

*where*

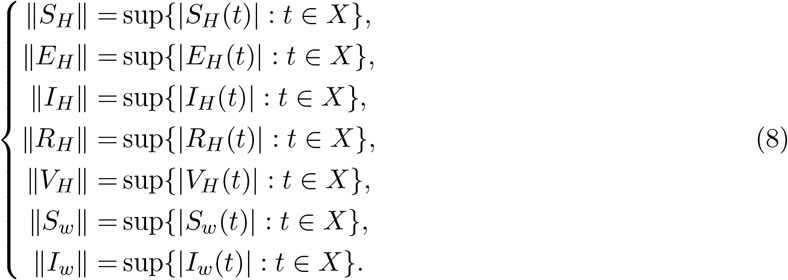

*Proof*. Applying the fractional ABC operator on both side of the equation (7) yields

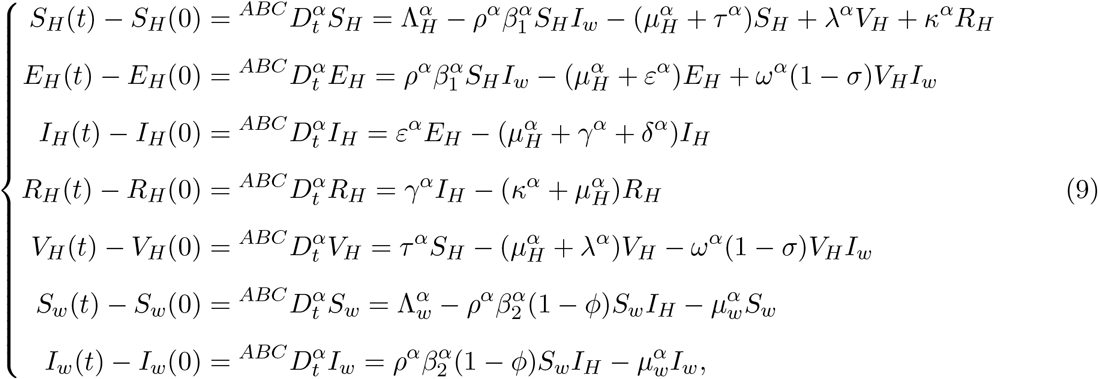

with the initial conditions *S*_*H*_(0) *>* 0, *V*_*H*_(0) ≥ 0, *E*_*H*_(0) ≥ 0, *I*_*H*_(0) ≥ 0, *R*_*H*_(0) ≥ 0, *S*_*w*_(0) ≥ 0 and *I*_*w*_(0) ≥ 0 where 0 ≤ *t <* ∞ and 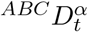 denotes the Atangana-Baleanu Caputo fractional derivative of order *α* ∈ (0, 1]. Applying Definition (2) on the model system Eq (6) we obtain;

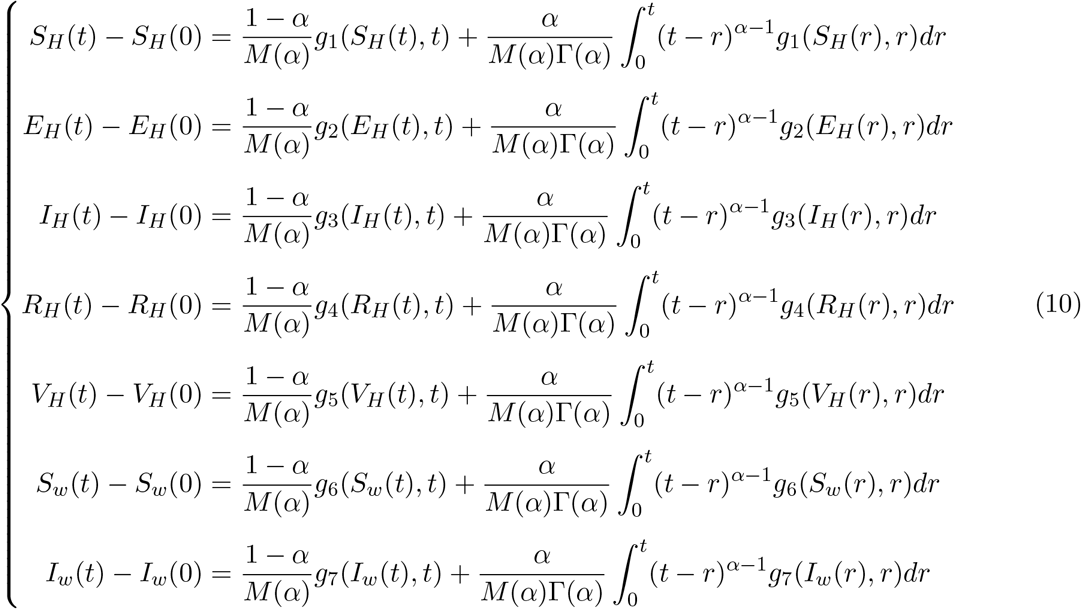

where

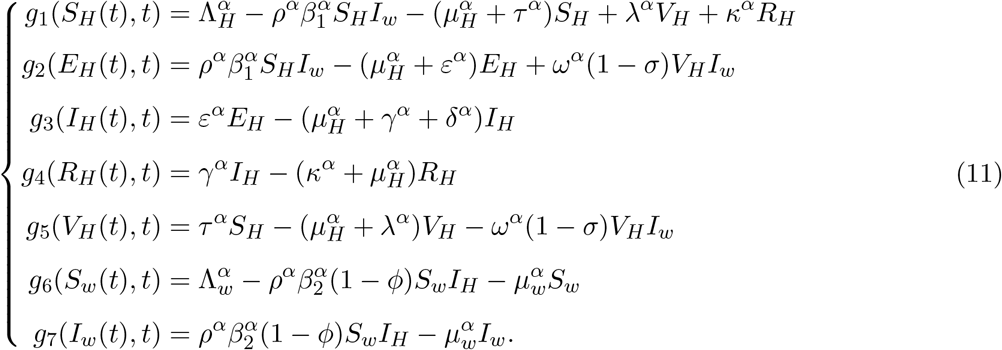

Given that *S*_*H*_(*t*), *E*_*H*_(*t*), *I*_*H*_(*t*), *R*_*H*_(*t*), *V*_*H*_(*t*), *S*_*w*_(*t*) and *I*_*w*_(*t*) have an upper bound, then *g*_1_(*S*_*H*_(*t*), *t*), *g*_2_(*E*_*H*_(*t*), *t*), *g*_3_(*I*_*H*_(*t*), *t*), *g*_4_(*R*_*H*_(*t*), *t*), *g*_5_(*V*_*H*_(*t*), *t*), *g*_6_(*S*_*w*_(*t*), *t*), and *g*_7_(*I*_*w*_(*t*), *t*) are said to satisfy the Lipschitz condition. Let *S*_*H*_(*t*) be two functions, such that

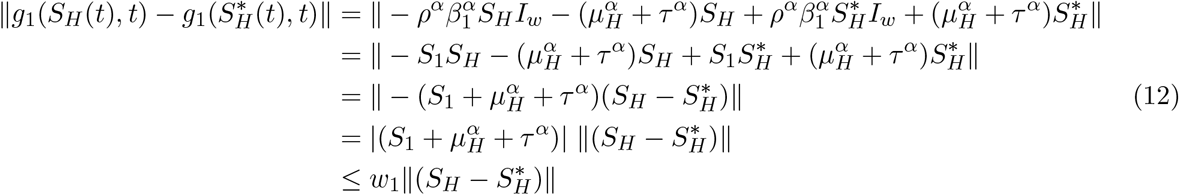

In the same procedure we obtain;

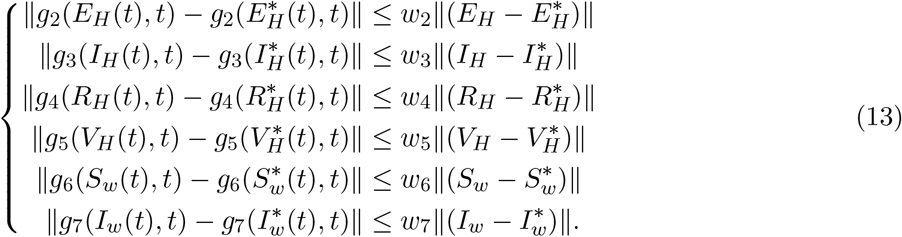

Hence, *w*_*i*_, *i* = {1, …, 7} are the corresponding Lipschitz constants that satisfies the Lipschitz condition for all the functions *S*_*H*_(*t*), *E*_*H*_(*t*), *I*_*H*_(*t*), *R*_*H*_(*t*), *V*_*H*_(*t*), *S*_*w*_(*t*) and *I*_*w*_(*t*). We can rewrite equation (10) recursively as

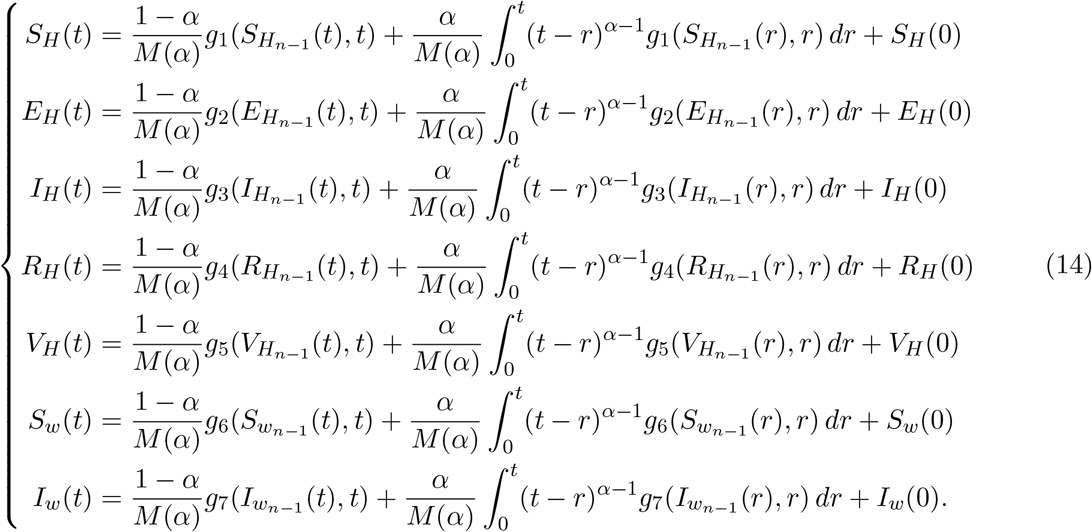

Taking the difference of the successive terms together with the initial conditions in equation (7), the following system of equations are derived, given by

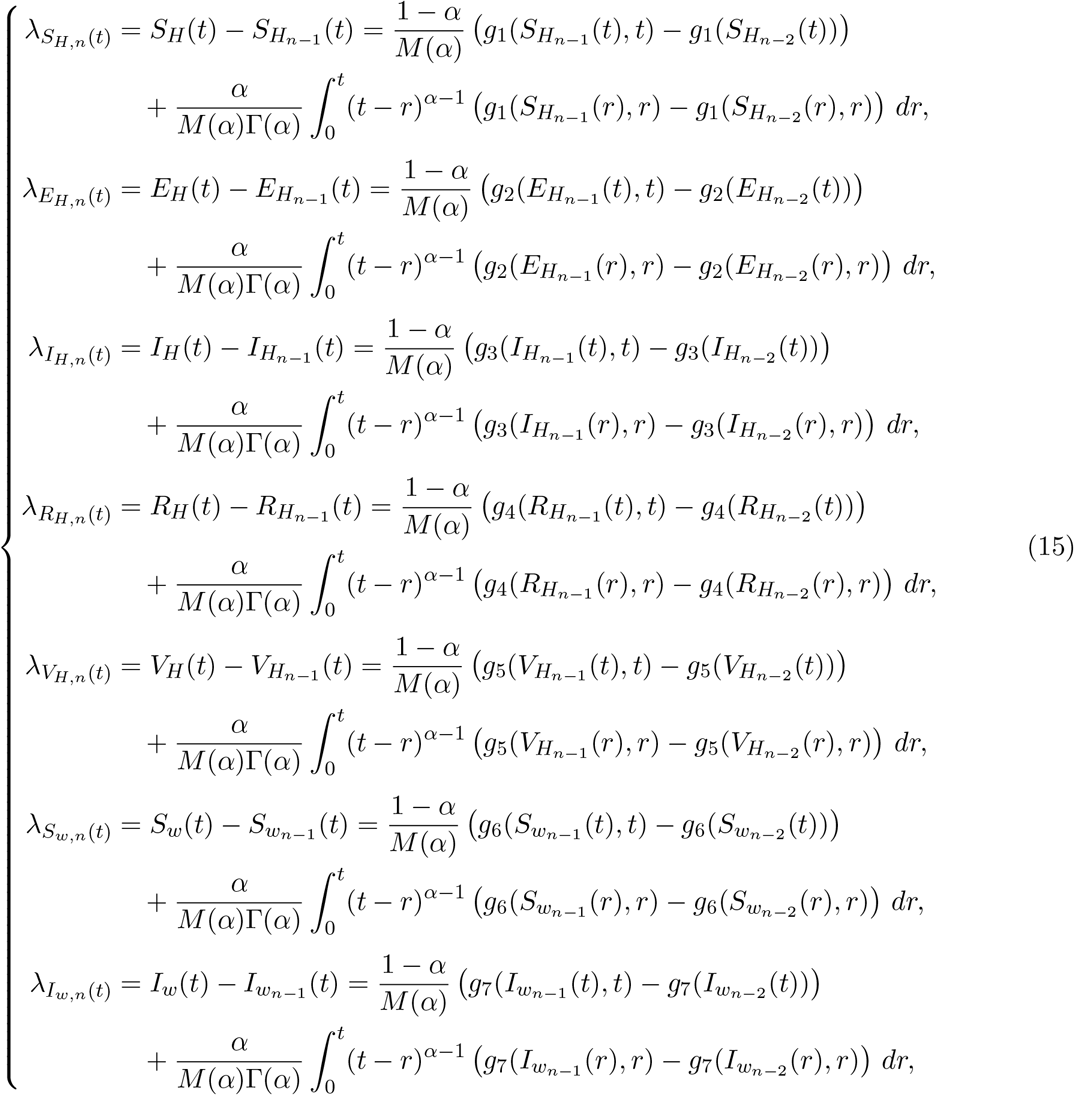

Also, it can be observed that

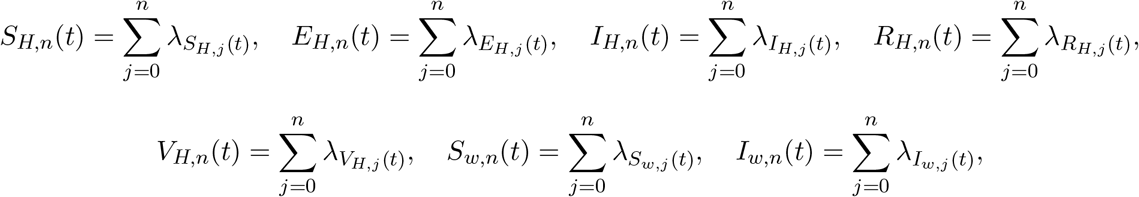

Using equations (12) and (13) and taking into account that

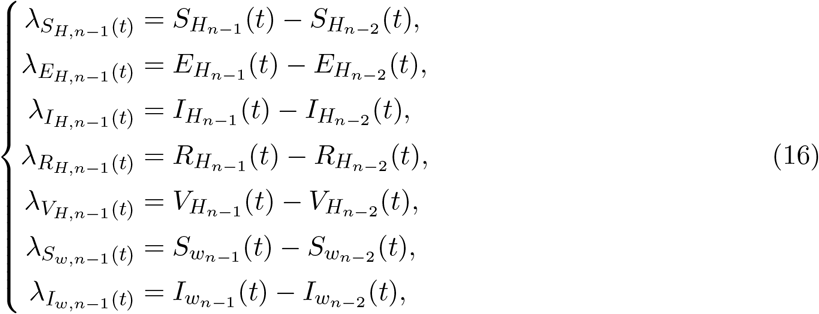

Then, the following are derived

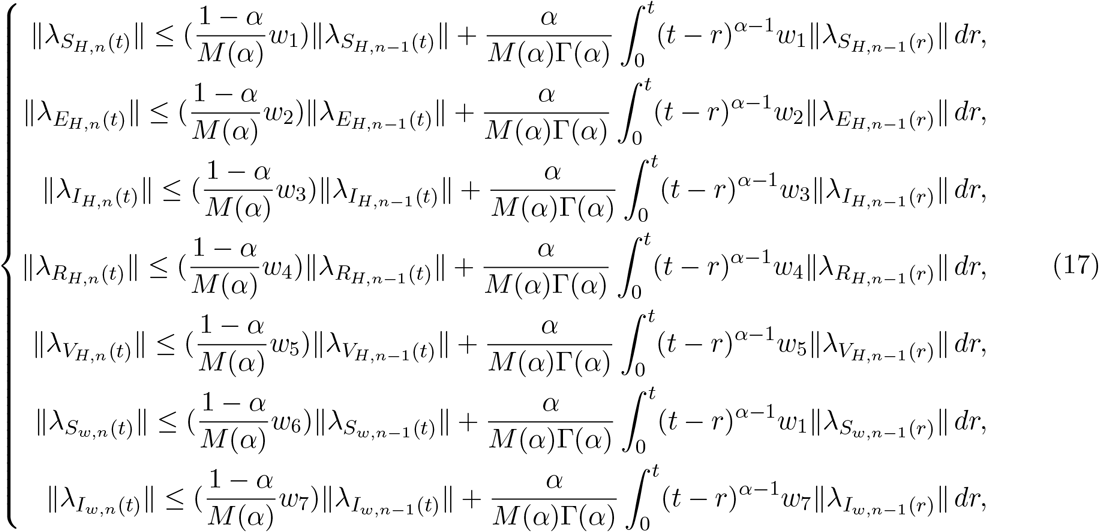

**Theorem 4.3**. *[41] The proposed fractional order Buruli Ulcer ABC operator model equation* (7) *possesses a unique solution for some t*_0_ ∈ [0, *T* ] *if the following condition holds true*.

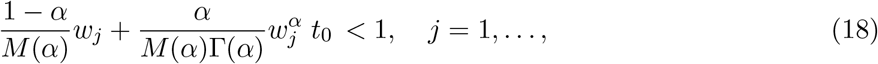

*Proof*. It is evident that *S*_*H*_(*t*), *E*_*H*_(*t*), *I*_*H*_(*t*), *R*_*H*_(*t*), *V*_*H*_(*t*), *S*_*w*_(*t*) and *I*_*w*_(*t*) are bounded functions and adhere to the Lipschitz condition. Furthermore, the functions *g*_1_, *g*_2_, *g*_3_, *g*_4_, *g*_5_, *g*_6_ and *g*_7_ also comply with the Lipschitz condition as demonstrated in(12) and (13). Therefore, by applying the recursive principle and using equation (17), the following system can be derived.

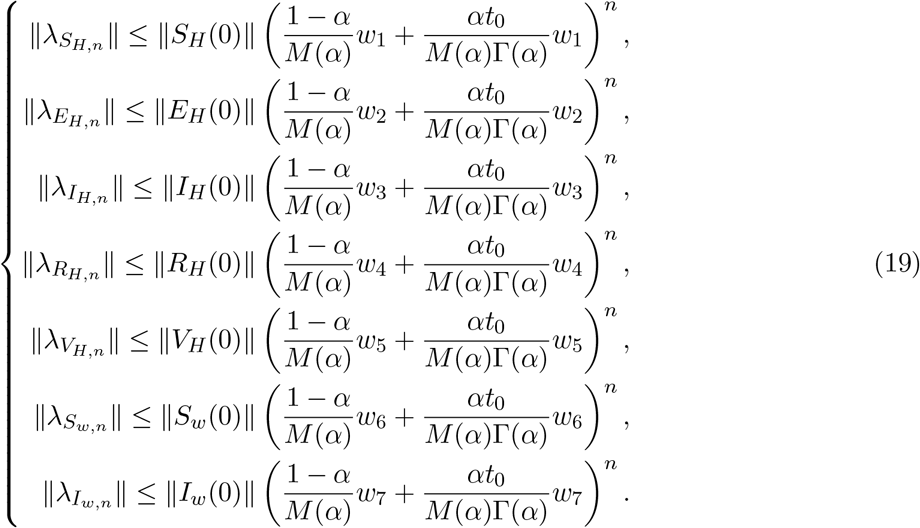

Thus, the sequences derived above exist and satisfy

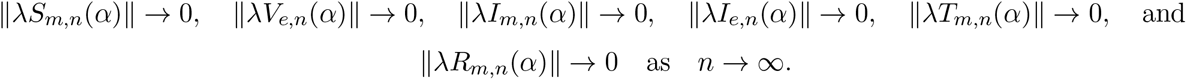

Additionally, from equation (19) and utilizing the triangular inequality for any *r*, we obtain

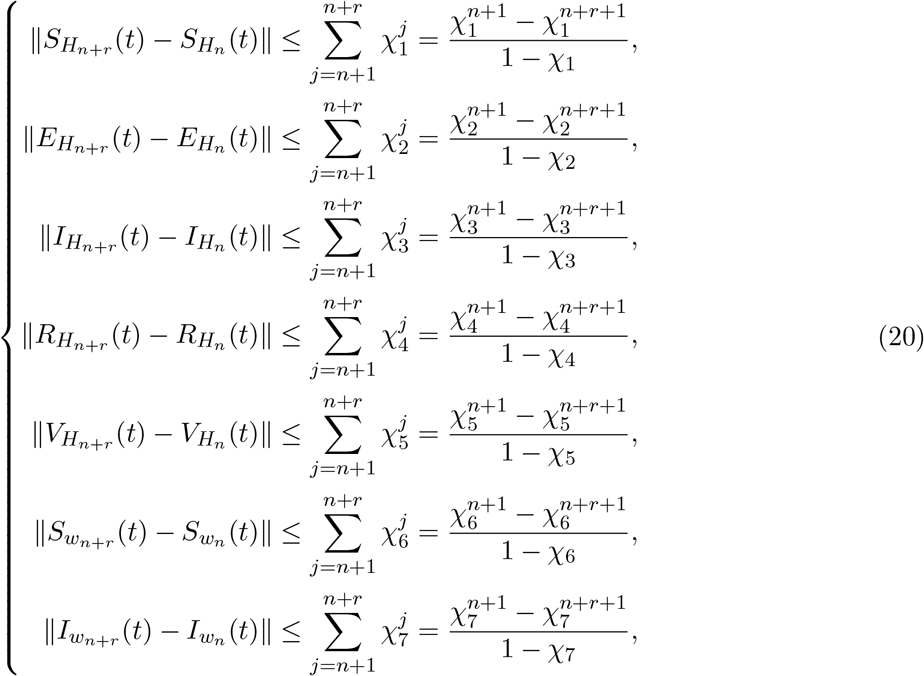

Where, 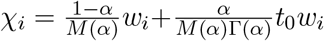 for *i* = {1, 2, …, 7}. As a result, *S*_*H,n*_, *E*_*H,n*_, *I*_*H,n*_, *R*_*H,n*_, *V*_*H,n*_, *S*_*w,n*_ and *I*_*w,n*_ form Cauchy sequences within *F* (*x*), converging uniformly. The limit of these sequences represents the unique solution to (7), demonstrated through the application of the limit theory in equation (14) as *n* approaches infinity. Thus, the existence of the unique solution for the fractional order ABC model system equation(7) is established.

## 5 Numerical Results

In this section, we present the numerical simulation of the fractional model (7). We consider the parameter values in (2) and observe the dynamics of both class of human and the water-bug population. We considered the following initial conditions *S*_*H*_(0) = 280,*E*_*H*_ = 80, *I*_*H*_(0) = 25, *R*_*H*_(0) = 20, *V*_*H*_(0) = 20, *S*_*w*_(0) = 700, *I*_*w*_(0) = 44. Varying orders of the fractional derivatives *α* = {0.8, 0.85, 0.9, 0.95} were used for the simulations.

From Figure (2), we observe that the number of susceptible humans *S*_*H*_ decline as they become exposed to the Buruli ulcer disease, while the number of infected individuals increase. A similar behaviour is seen in the water-bugs. The number of infected water-bugs increases whiles the susceptible water-bugs decrease. This trend illustrates the natural progression of the Buruli ulcer disease outbreak, where the susceptible population diminishes as more individuals become infected. Also, we observe that both the vaccinated and recovered humans decrease. In the Figures 2a to 2f, we plot the solution trajectory for each compartment with varying fractional orders *α* = {0.8, 0.85, 0.9, 0.95 }.

**Figure 1:**
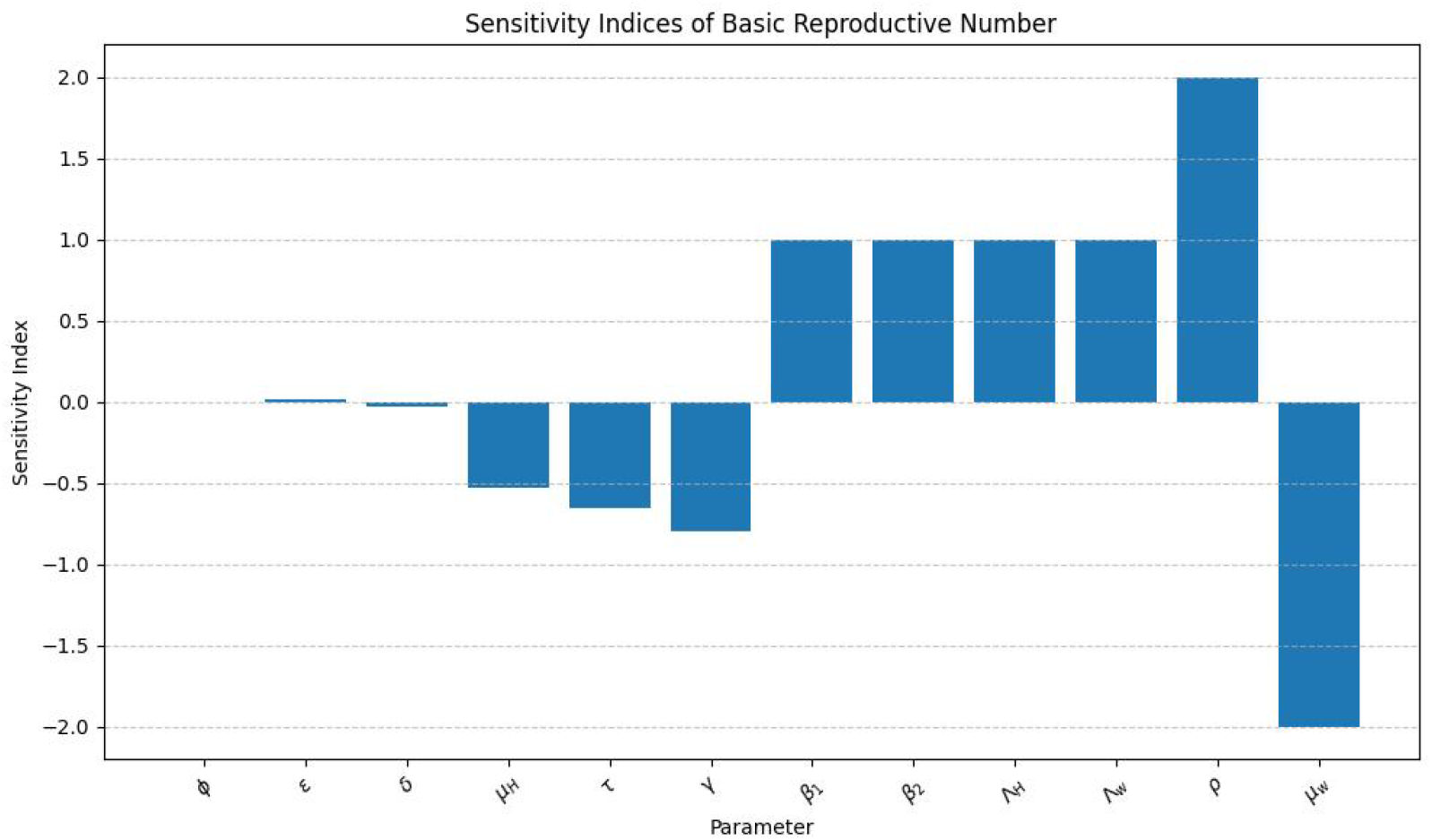
Sensitivity indices of the basic reproductive number.

**Figure 2:**
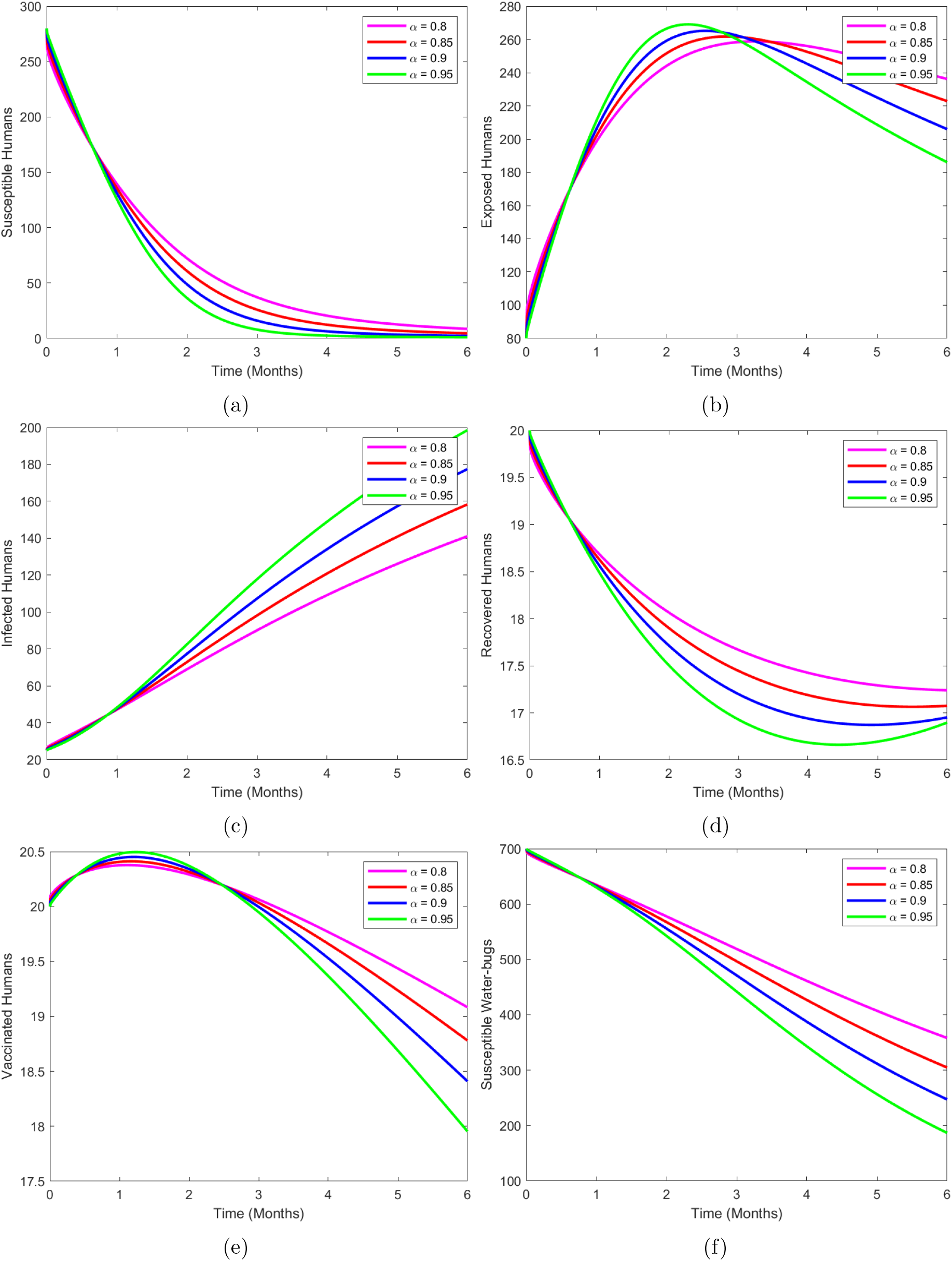
Simulation of the fractional model model without control.

**Figure 3:**
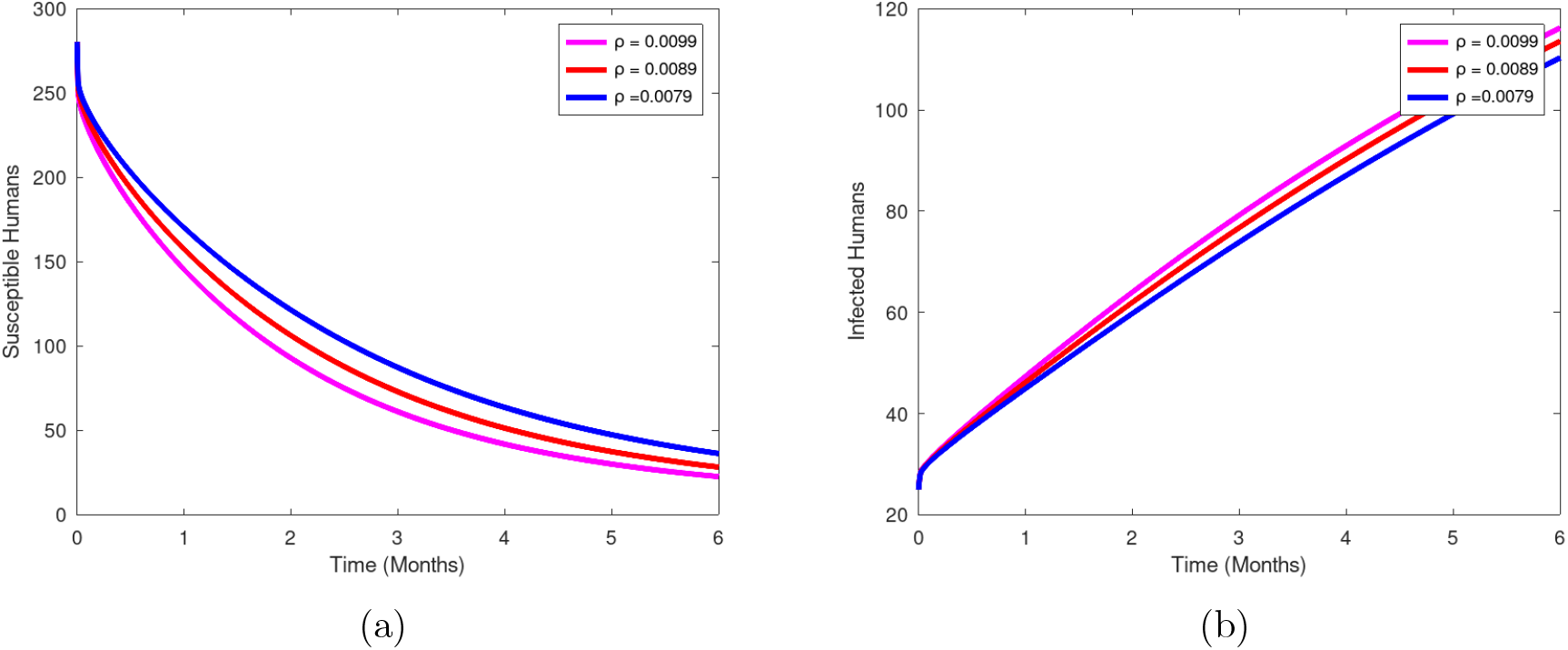
The dynamics of Susceptible and infected humans when the parameter *ρ* is varied.

The graph (3a) and (3b) shows the dynamics of the susceptible and infected human class with varying parameter *ρ*. It can be observed that decrease (increase) in the biting rate of the water bugs on humans, *ρ* lead to an increase (decrease) in the susceptible humans and a decrease (increase) in the infected humans. An increase in the recovery rate *γ* from figure (4) leads to a rise in the number of susceptible humans and a decrease in the infected humans class. From Figure (5) we observe that when the parameter *τ* is varied there is much effect on the susceptible and vaccinated human class. When the value of the parameter *τ* is increased the vaccination of humans rises as the susceptible humans decline.

**Figure 4:**
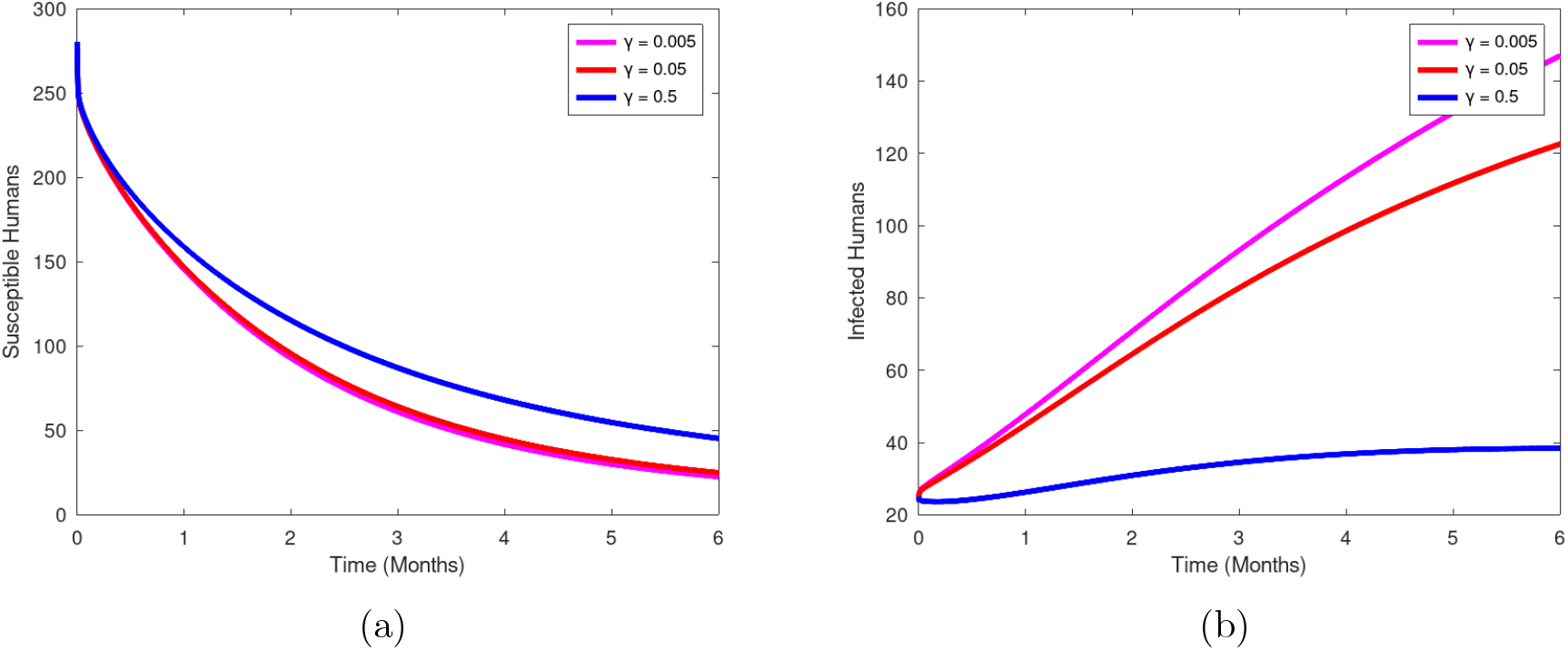
The dynamics of susceptible and infected humans when the parameter *γ* is varied.

**Figure 5:**
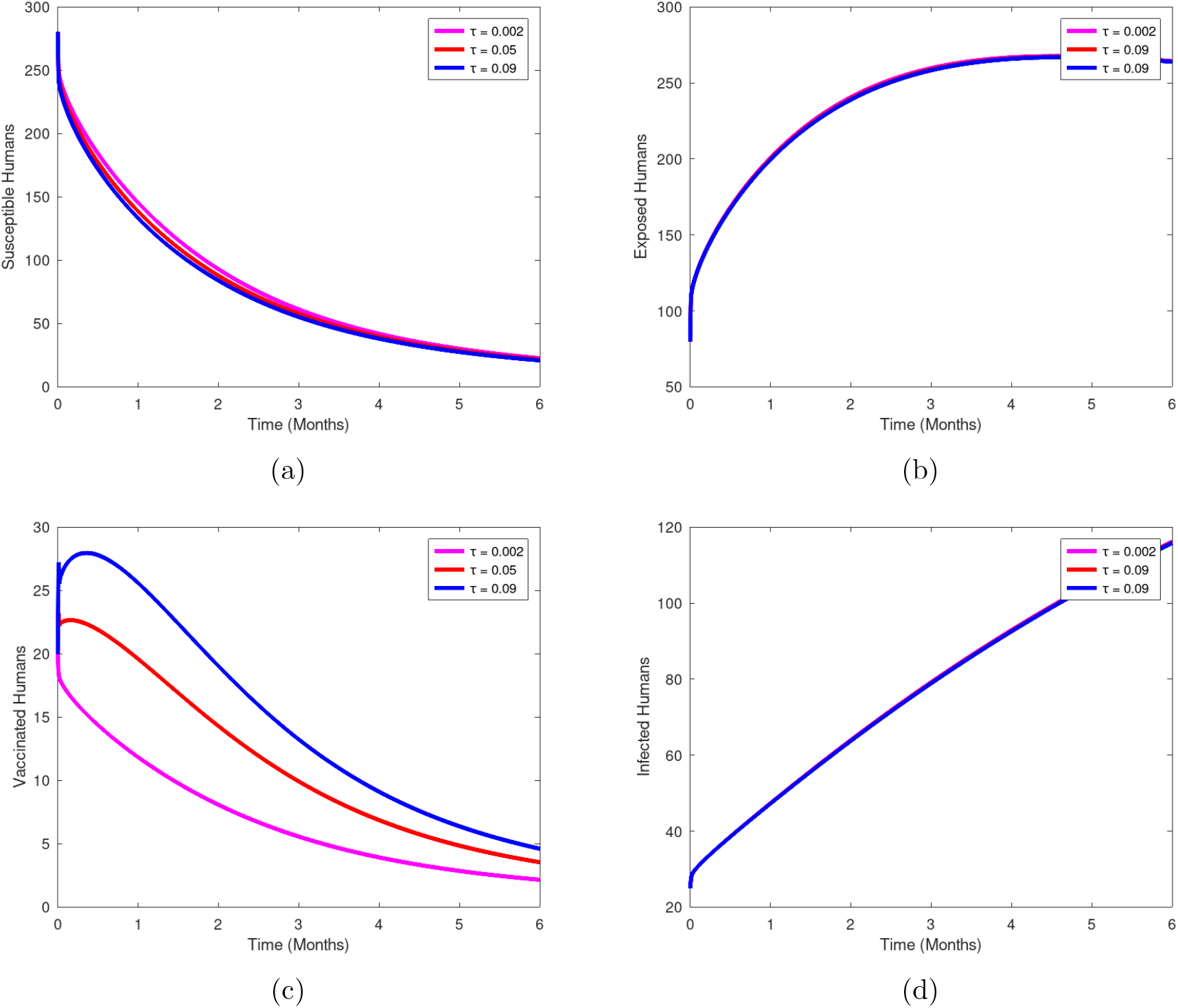
The dynamics of the Buruli model when the parameter τ is varied.

**Figure 6:**
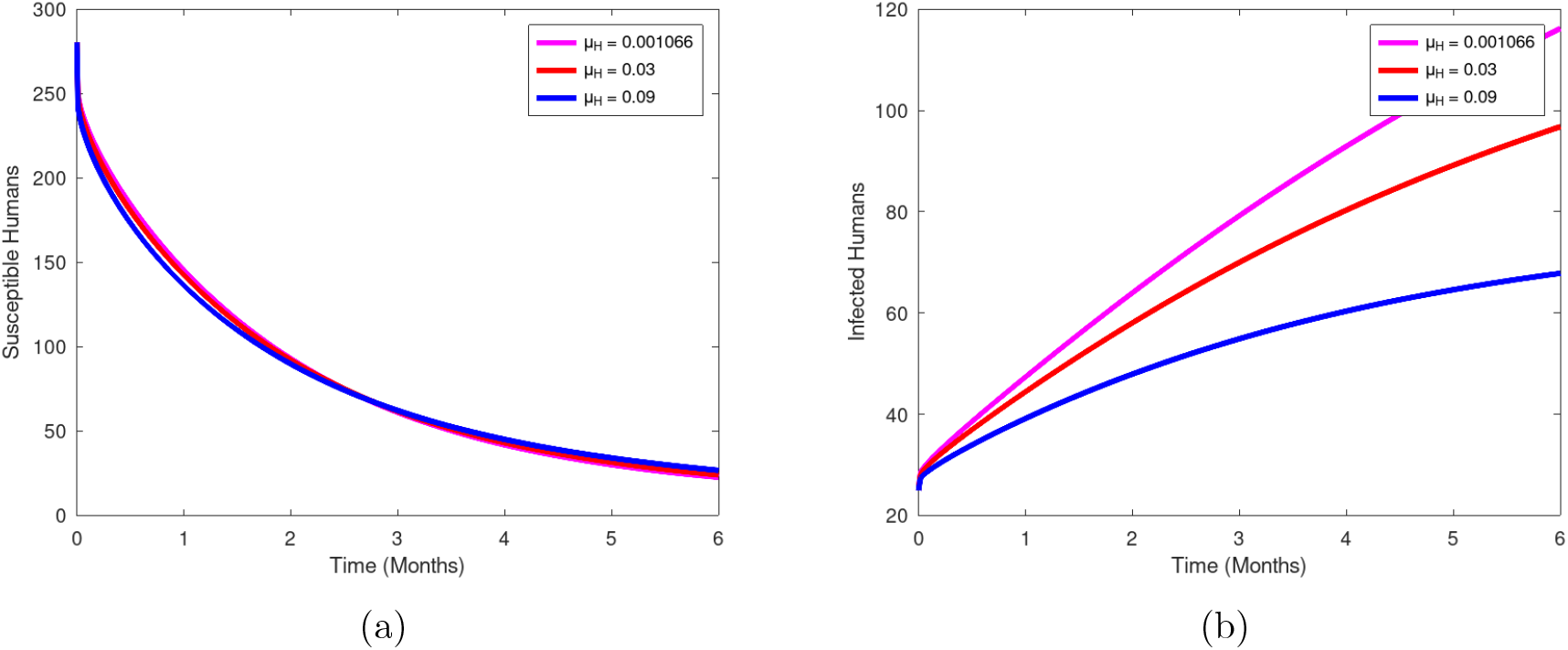
The dynamics of Susceptible and infected humans when the parameter *µ*_*H*_ is varied.

**Figure 7:**
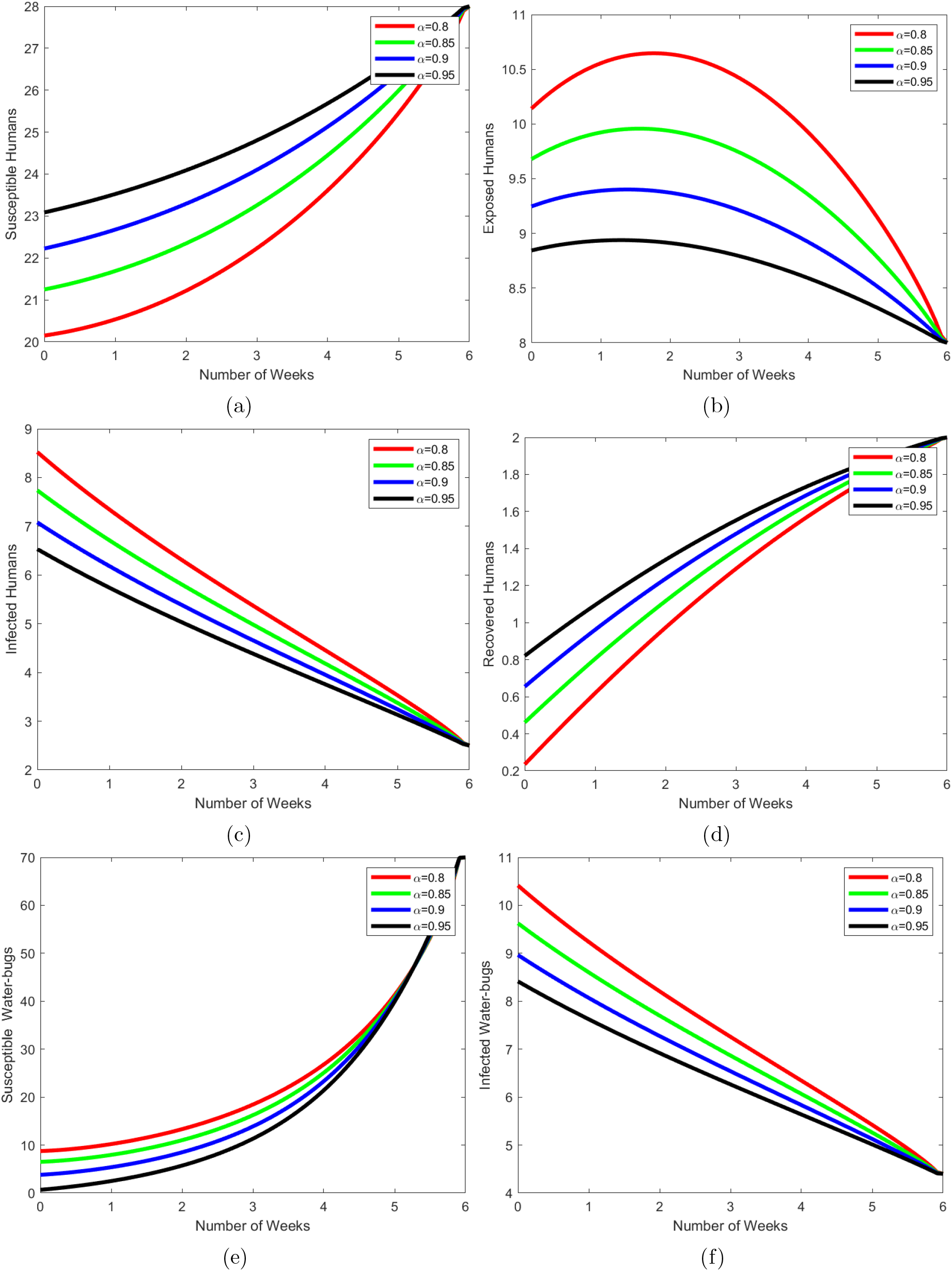
Applying all control strategies

**Figure 8:**
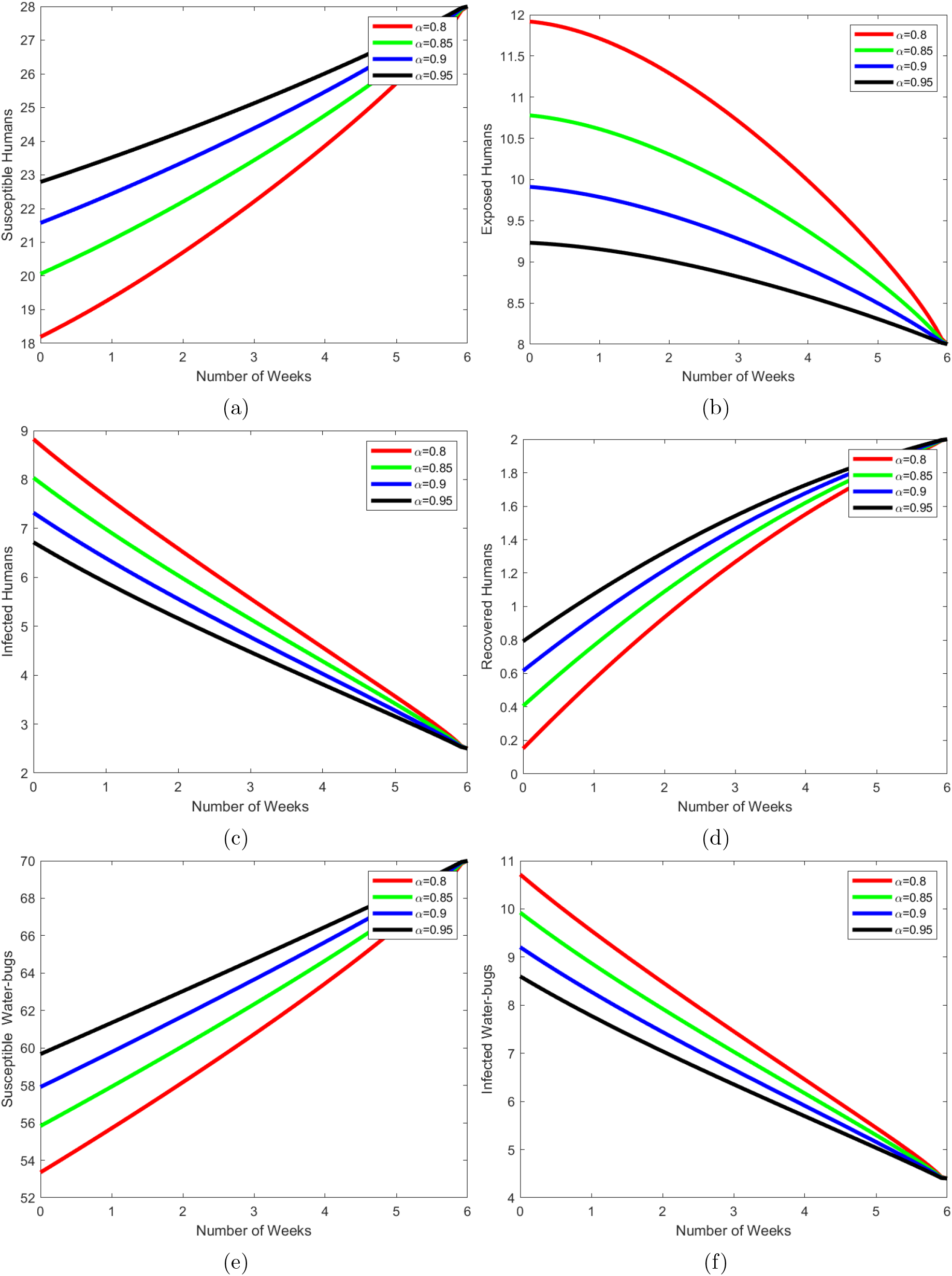
The intervention by education on wearing protective clothing, rate of vaccination and treatment of infected humans.

**Figure 9:**
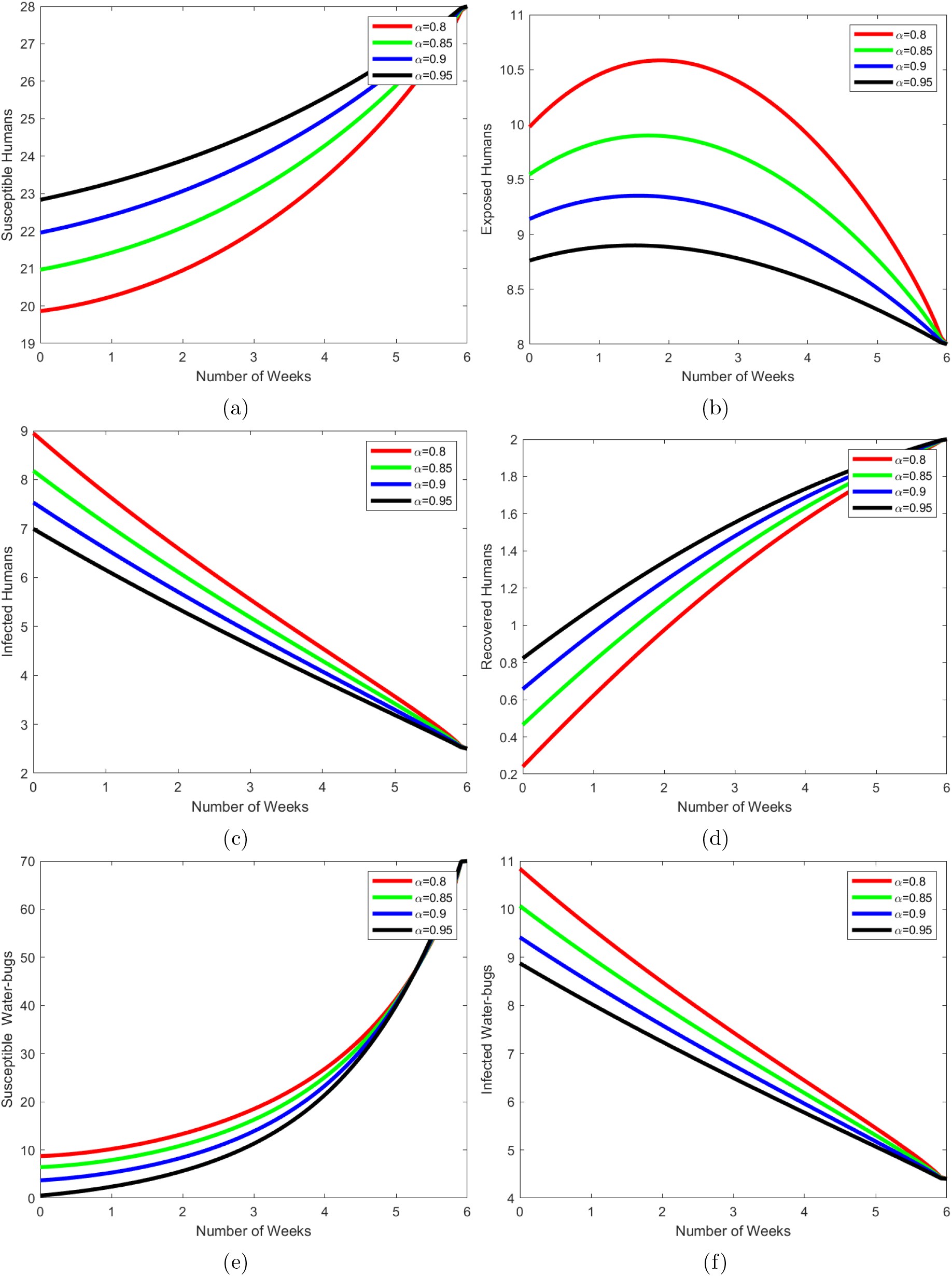
The intervention by education on wearing protective clothing, rate of vaccination and applying insecticides on water-bug.

**Figure 10:**
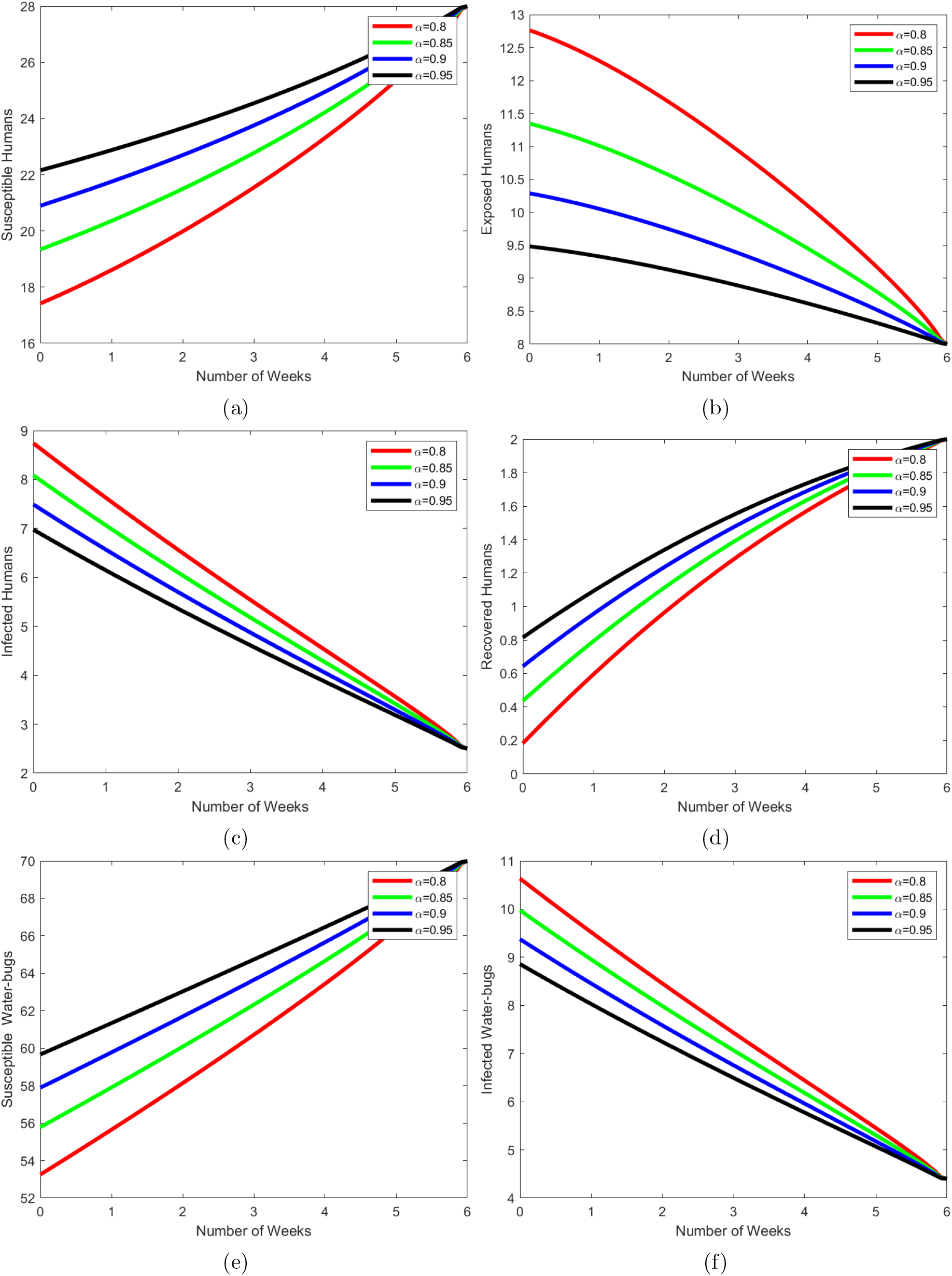
The intervention by education on wearing protective clothing and rate of vaccination only.

## 6 Fractional Optimal Control of Buruli Ulcer

In this section, we present the Buruli ulcer fractional optimal control problem. From the model (7) we modify it by applying the the optimal control interventions for effective management of the Buruli ulcer infection. The control interventions arose as a result of the computation of the sensitivity indices. We incorporate in the new model four control interventions namely; health education *u*_1_(*t*), vaccination rate *u*_2_(*t*), effective treatment of infected humans *u*_3_(*t*), and spraying insecticides on waterbugs population *u*_4_(*t*). The goal is to minimize the number of exposed humans *E*_*H*_, infected humans *I*_*H*_ and the population of water-bugs *N*_*w*_ while minimizing the cost of control interventions. Hence the objective function *J*(*u*) can be formulated as

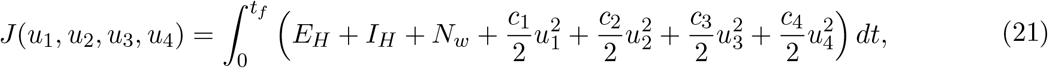

subject to the fractional optimal control problem with interventions,

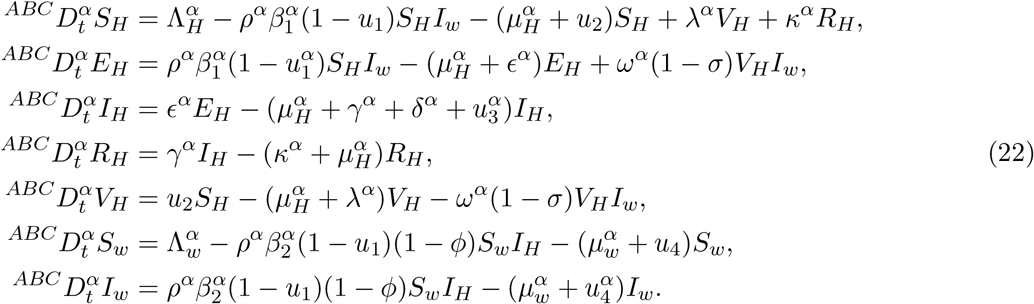

with positive initial conditions :

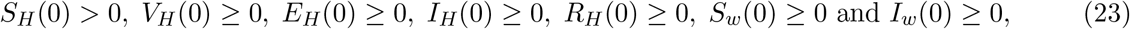

where *c*_1_, *c*_2_, *c*_3_, *c*_4_ are the weights associated with the cost of the control measures. The primary goal of the fractional optimal control problem is to identify an optimal control 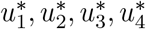 such that

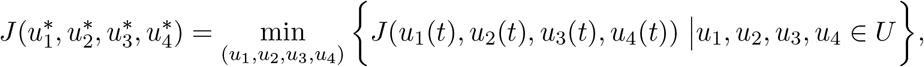

where the admissible set of controls is given by

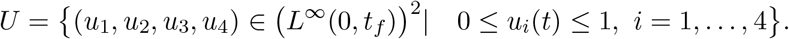

### 6.1 Characterization of Optimal Control

The existence outcome for optimal control from the adjoint variable of the state variables satisfies the following set of differential equations, and the required criteria that an optimal control must meet were obtained from Pontryagin’s Maximum Principle according to [39, 45]. With regard to the controls *u*_1_, *u*_2_, *u*_3_, *u*_4_, this principle transforms system (22) into a problem of minimizing a Hamiltonian *H* point-wise. Our initial step will be finding the Lagrangian and Hamiltonian for the optimal control problem.

The Lagrangian formulation is given by

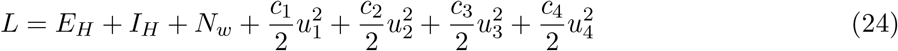

The Hamiltonian associated with the control problem is

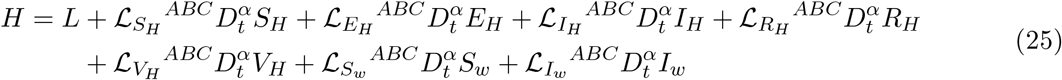

Hence by Pontryagin’s Maximum Principle, the Hamiltonian *H* is as follows:

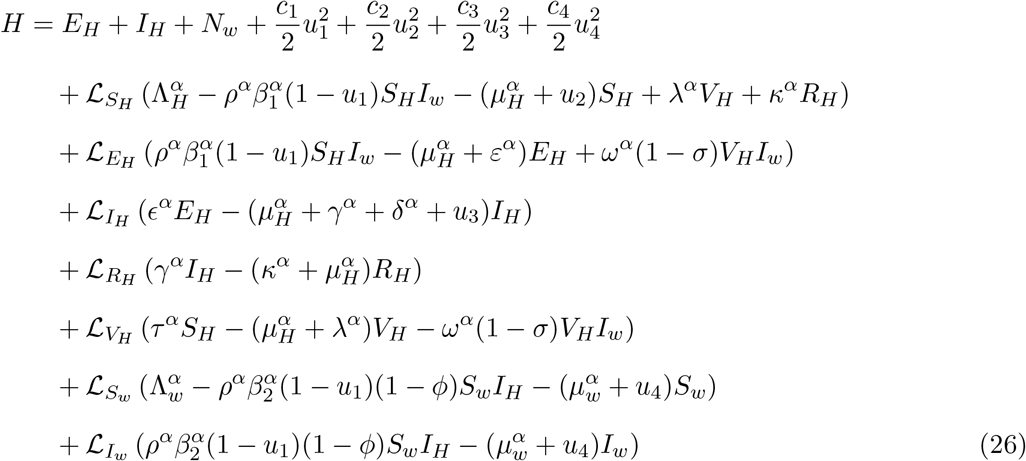

where 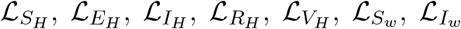 are the adjoint variables or the Lagrangian multipliers. The maximal principle of Pontryagin states that if (X,U) gives an optimal solution to an optimal control problem, then there exists a nontrivial vector function 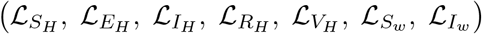 with the following properties.

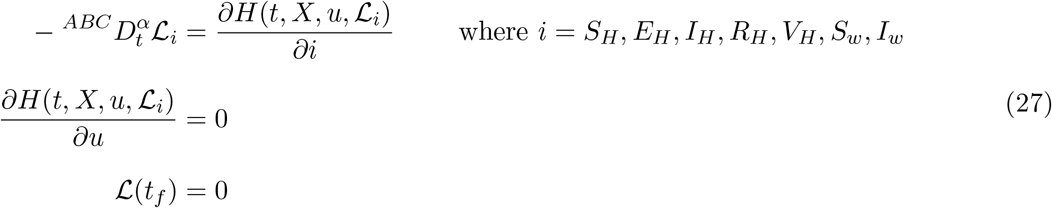

where *X* and *U* are the optimal solution and controls respectively. The existence of the optimal control 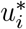 can be establish in the following theorem.

**Theorem 6.2**. *Let* 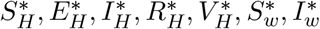 *be the associated solution to the following optimal control problem and* 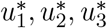 *and* 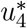*be the optimal control that minimizes J*(*u*_1_, *u*_2_, *u*_3_*u*_4_) *over U. Then there exist adjoint functions* ℒ_*i*_ *satisfying the following three results*

1. Equations of adjoint state variables

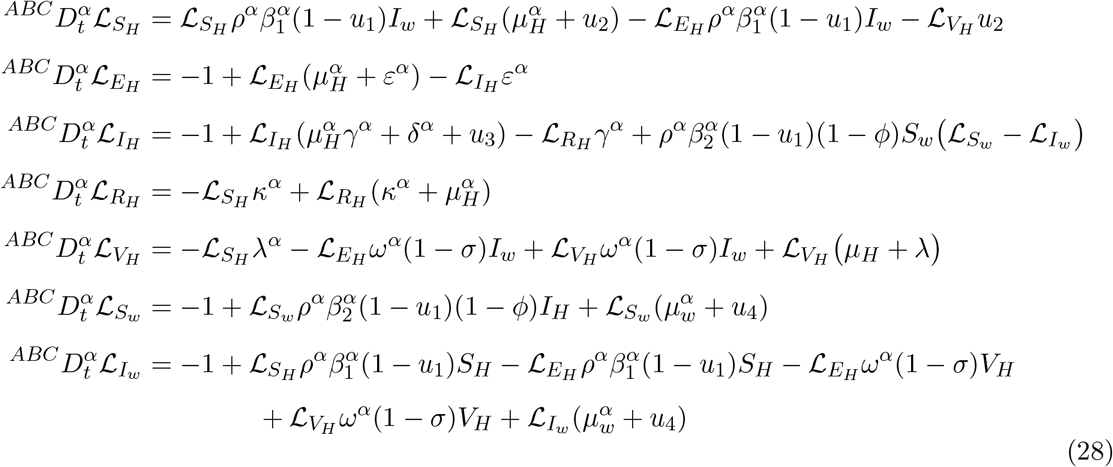

2. With transversality conditions

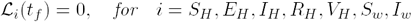

3. Furthermore, the optimality condition for FOCP as follows:

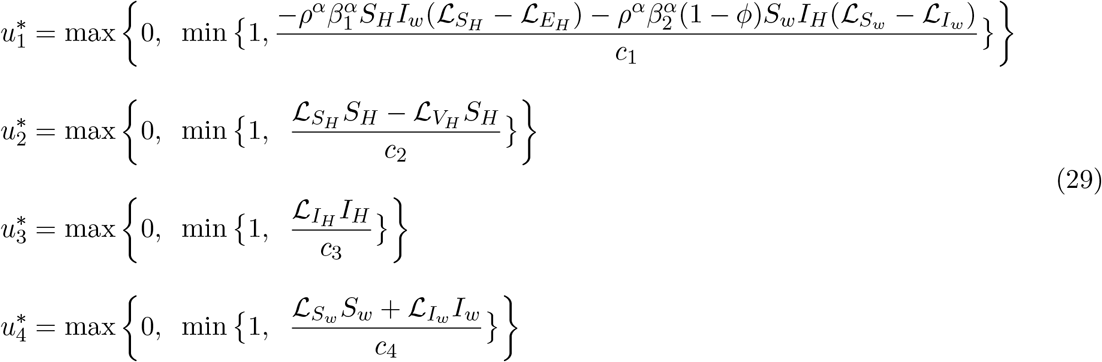

*Proof*. In trying to show the proof to the above theorem we use the Hamiltonian function (25) to get the adjoint and transversality criteria. We compute the adjoint system by employing Pontyagin’s maximal principle as follows;

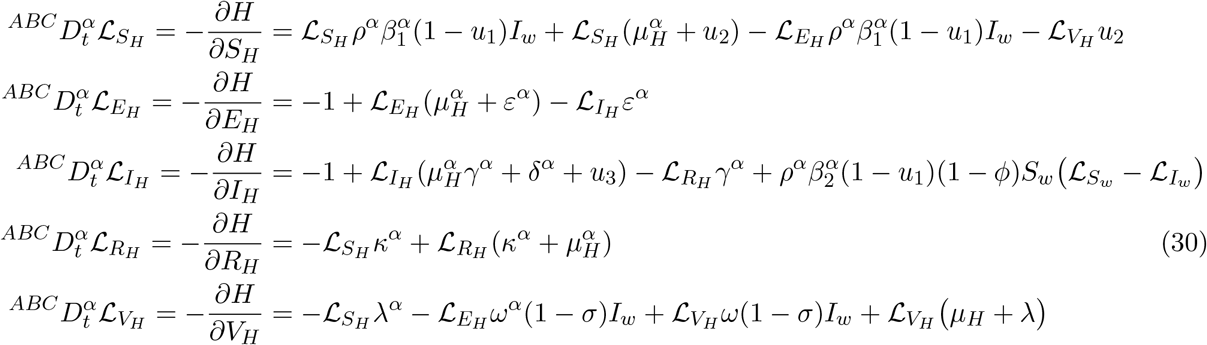

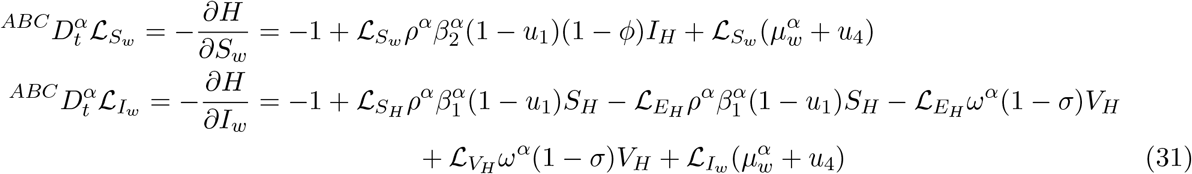

with transversality condition

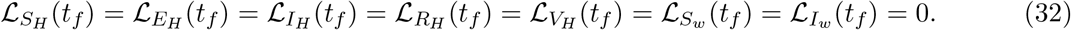

Also the optimal functions 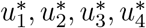 satisfies

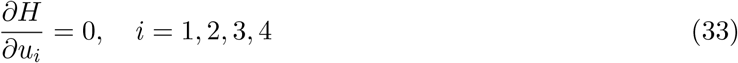

Hence by making use of (33) the optimal control variables are obtained as

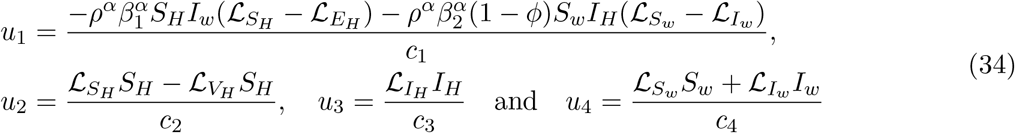

Hence the proof is complete.

The uniqueness of the optimality system (28) was obtained as a result of the priori boundedness of the state system (22), the adjoint system, and so on. To ensure the uniqueness of the optimality system, we limit the duration of the time interval [1, *t*_*f*_ ]. The optimal state can be found by substituting 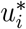 into (22).

## 7 Numerical Simulations

In this section, we look at how the interventions affect the Buruli ulcer transmission in a population. We used the modified PECE method of Adam-Bashforth Moulton to solve the adjoint variable (28) and the fractional optimal control problem numerically, see, e.g. [39]. In order to reduce the incidence of Buruli ulcer infections in humans and water bugs, respectively, we use the weight *c*_1_, *c*_2_, *c*_3_, *c*_4_, and parameter values in table (2). We vary the fractional order for four *α* values and set the time limit at six months. There are many combinations of intervention strategies we could consider however we limit ourselves to the following intervention strategies implemented in the simulations shown in the graphs below.

**Table 2:**
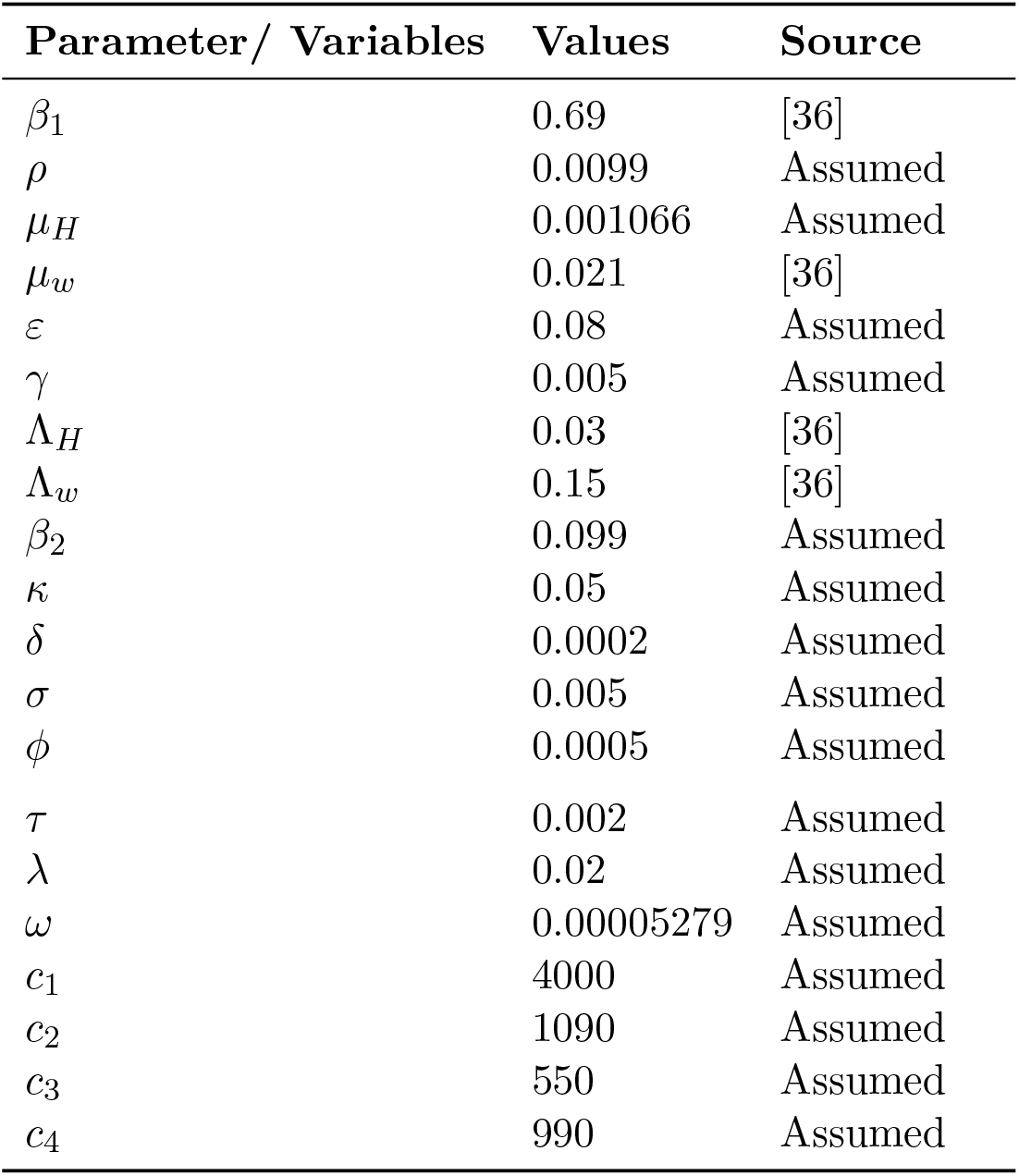
Model’s parameter values.

### 7.1 Strategy A: All strategies

The intervention method here present the solution of the optimal control system (22) when we focus on all the control variables that is *u*_1_ = *u*_2_ = *u*_3_ ≠ *u*_4_ 0. From the figures in (7a)-(7f) we observe that there is a rise in the susceptible humans and a decrease in the number of infected humans due to the decline in the number of people exposed to the disease. This then leads to high recovery rate. Concurrently there is a slow increase in susceptible water-bugs as the infected water-bugs decreases.

### 7.2 Strategy B: (*u*_1_), (*u*_2_), and (*u*_3_) only

In this strategy, we consider the case where education on the use of protective clothing, the rate of vaccination and treatment of infected humans are implemented, when *u*_1_ = *u*_2_ = *u*_3_ ≠ 0, *u*_4_ = 0. From the figures in (8a)-(8f), we observe that the populations of the susceptible and recovered humans increases as the infected humans decline relative to the uncontrolled situation in figure (2). This is as a result of a rapid decline in the number of humans exposed to the disease in the endemic region.

In the same vane we observe a decreasing effect on the infected water-bugs and a linear rise in the susceptible water-bugs.

### 7.3 Strategy C: (*u*_1_), (*u*_2_), and (*u*_4_) only

The intervention method present here looks similar to strategy A where there is a rise in the susceptible and recovered humans as a result of decline in exposed humans which leads to a rapid decrease in the infected human. Simultaneously there is a rapid decline in the infected water-bugs and slowly increase in the susceptible water-bugs.

### 7.4 Strategy D: (*u*_1_) and (*u*_2_) only

The strategy shown over here is a combination of education on wearing protective clothing and rate of of vaccination where *u*_1_ = *u*_2_ ≠ 0, *u*_3_ = *u*_4_ = 0. It is clearly observed that this strategy is also very effective as the rate at which people are exposed to the disease falls which leads to increase in the number of susceptible and recovered humans and a decrease in the infected humans. There is also rise in the susceptible water-bugs and a fall in infected water-bugs respectively.

### 7.5 Strategy E: (*u*_3_) and (*u*_4_) Only

The intervention method here presents the solution of the optimal control system (22) when we focus on treating infected humans and applying insecticides on the water-bug population only, that is *u*_1_ = *u*_2_ = 0, *u*_3_ = *u*_4_ ≠ 0. From the figures (11a)-(11), combining these two control interventions leads to a rapid increase in the susceptible humans and decline in the number of infected humans as there is a decrease in the number of people exposed to outbreak of the disease. We also observe that recovered humans increases whiles there is rapid decline in the infected water-bugs and slowly increase in the susceptible water-bugs.

**Figure 11:**
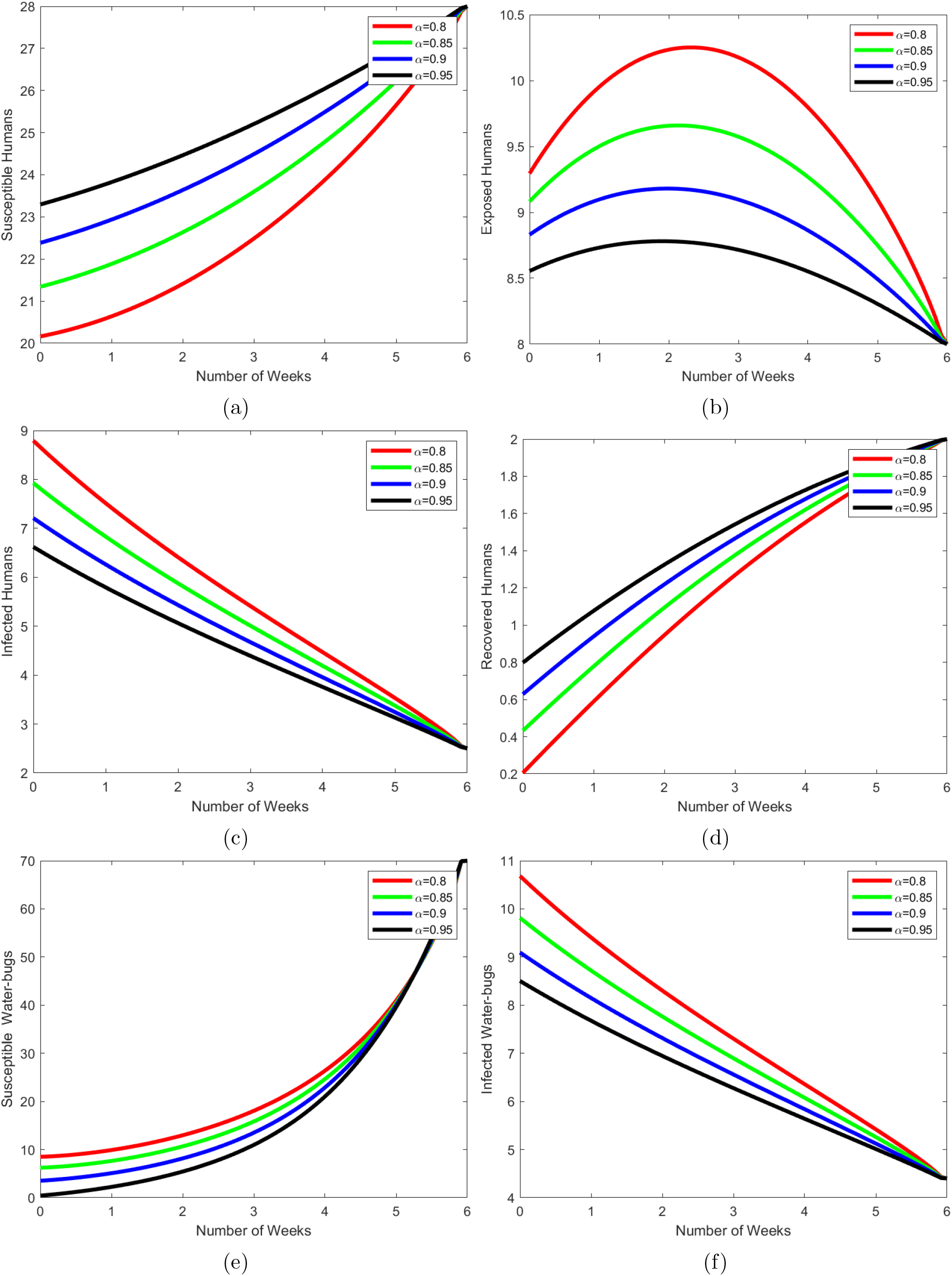
Intervention by treating infected humans and Spraying insecticides on Water-bugs only.

## 8 Conclusion

In this study, we developed a fractional optimal control model for Buruli ulcer transmission that includes health education on the use of protective clothes, vaccination rates, treatment of infected persons, and insecticide spraying on the water bug population. We used ABC fractional order derivatives to test the effect of fractional order derivatives. The basic features of the model without control variables were examined, revealing that the model is both biologically and mathematically well-posed. We subsequently formulated the fractional optimal control problem by using Pontryagin’s Maximum Principle, the optimal control problem was solved. We then presented a numerical simulation of the fractional model without control and with control. Several control strategies were implemented and we observed from the graphs that when the control measures are applied relative to the one without control there is an increase in the susceptible, recovered, and vaccinated humans while there is a decline in infected humans and infected water-bugs. We then conclude and recommend that governments in the endemic regions should invest more in finding perfect vaccines, implement comprehensive health education on wearing protective cloths, ensure early and effective treatment of infected humans, and applying insecticides on the vectors that carry the *Mycobacteria Ulcerans*.

## Data Availability

No Data associated in the manuscript

## Author Contribution

**SN**: Writing review & editing, Methodology, Conceptualization. **EA**: Writing - original draft, Simulations, Formal analysis. **MD**: Writing - review & editing. **SM**: Writing - review & editing, Conceptualization, Methodology

## Data Availability Statement

No Data associated in the manuscript

